# Continuous-Time and Dynamic Suicide Attempt Risk Prediction with Neural Ordinary Differential Equations

**DOI:** 10.1101/2024.02.25.24303343

**Authors:** Yi-han Sheu, Jaak Simm, Bo Wang, Hyunjoon Lee, Jordan W. Smoller

## Abstract

Suicide is one of the leading causes of death in the US, and the number of attributable deaths continues to increase. Risk of suicide-related behaviors (SRBs) is dynamic, and SRBs can occur across a continuum of time and locations. However, current SRB risk assessment methods, whether conducted by clinicians or through machine learning models, treat SRB risk as static and are confined to specific times and locations, such as following a hospital visit. Such a paradigm is unrealistic as SRB risk fluctuates and creates time gaps in the availability of risk scores. Here, we develop two closely related model classes, Event-GRU-ODE and Event-GRU-Discretized, that can predict the dynamic risk of events as a continuous trajectory based on Neural ODEs, an advanced AI model class for time series prediction. As such, these models can estimate changes in risk across the continuum of future time points, even without new observations, and can update these estimations as new data becomes available. We train and validate these models for SRB prediction using a large electronic health records database. Both models demonstrated high discrimination performance for SRB prediction (e.g., AUROC > 0.92 in the full, general cohort), serving as an initial step toward developing novel and comprehensive suicide prevention strategies based on dynamic changes in risk.

## INTRODUCTION

More than 700,000 people die by suicide worldwide annually, according to the World Health Organization^1^. In the US, suicide rates continue to increase despite calls for and efforts in suicide prevention^2^. One of the cornerstones of effective suicide prevention is identifying individuals at high risk for suicide-related behaviors (SRBs) to enable early intervention. Healthcare settings provide an important opportunity for risk assessment as most people who attempt or die by suicide are seen by a healthcare provider in the preceding weeks^3^. In recent years, the application of statistical and machine learning models to healthcare data have shown promise in improving prediction of suicide-related behavior^4–10^. Nevertheless, whether based on clinician evaluations alone or in conjunction with newer data-driven approaches^4,5^, current assessment methods are largely confined to specific time points and settings, such as following a healthcare visit, and treat risk as a static estimate over a given prediction window (Figure 1(a)). Such approaches are intrinsically limited given the dynamic nature of SRB risk, potentially compromising efforts to improve prevention. The precision of static risk estimates will decay with increasing intervals since the time point of prediction, progressively misaligning estimated and actual risk levels. In addition, in the absence of frequent healthcare visits, gaps in the availability of SRB risk estimates would occur, which could contain periods of high risk (i.e., the “uncovered time” depicted in Figure 1(a)). Ideally, an effective SRB risk estimate tool would provide a continuous-time, dynamic trajectory of risk that covers all future time points and can be updated with ongoing observations (Figure 1(b)).

**Figure 1.**
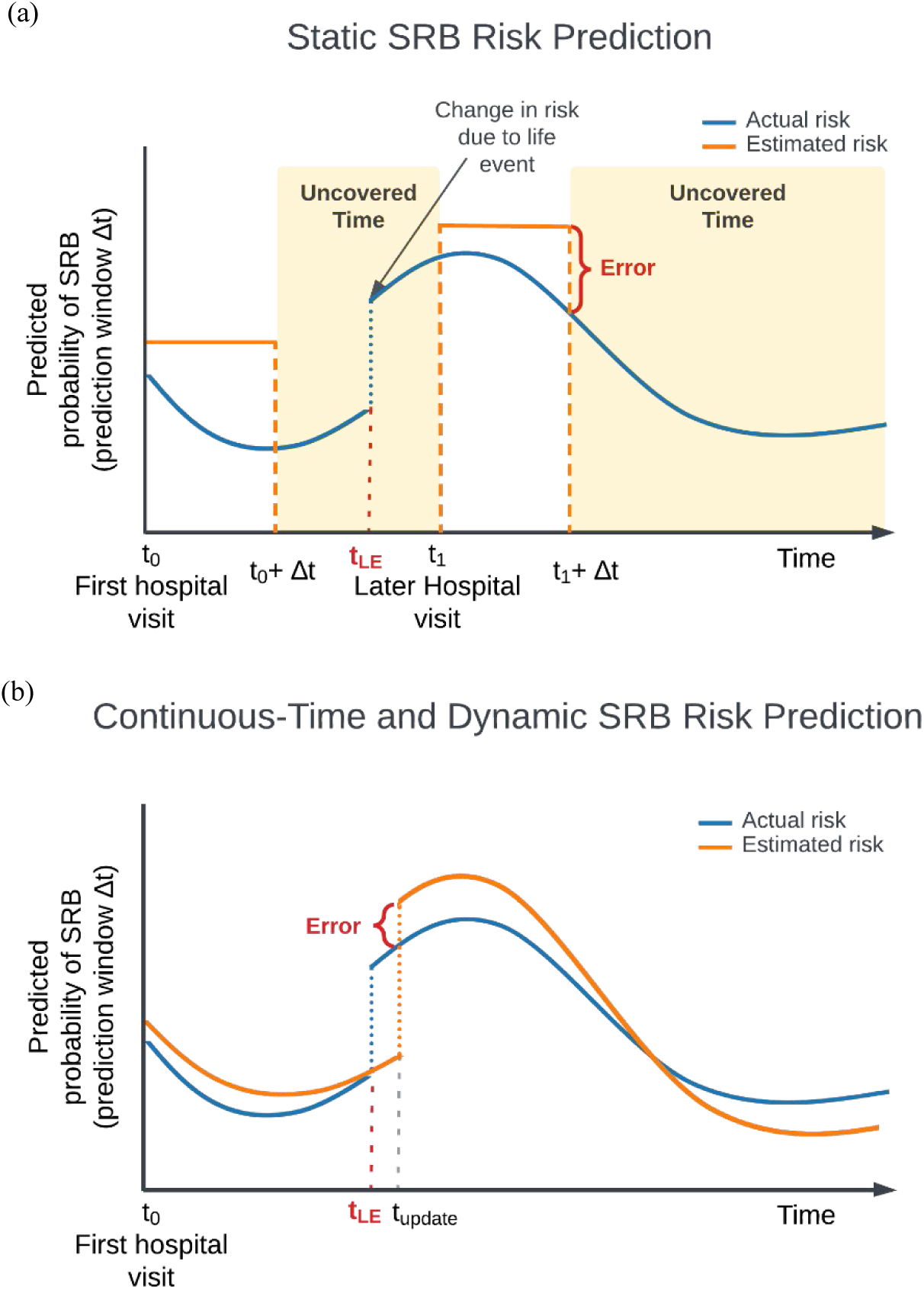
Schematic diagram for (a) static SRB risk predictions. In the static paradigm, risks of SRB are modeled as uniform probabilities for a pre-defined prediction window (Δt) evaluated at a specific time point, usually following a hospital visit (e.g., t_0,_ t_1_). Δt may vary in length based on the query. The estimated static risk is carried over through the entire prediction window to represent the SRB risk at any given point until the end of the prediction window (e.g., between t_0_ and t_0_+Δt). This approach is vulnerable to errors due to its inherent mismatch with the dynamic nature of SRBs, and fails to provide comprehensive coverage of estimated risk throughout the entire trajectory. On the x-axis: time since the first hospital visit; on the y-axis: actual (blue) and modeled (orange) SRB risk (cumulative incidence) for a given prediction window. t_0_: the time of the first hospital visit; t_1_: a subsequent hospital visit when new information regarding a life event is recorded; Δt_1_: the corresponding prediction window queried; Error: the discrepancies between estimated and actual risks; t_LE_: the time a life event occurred, potentially affecting SRB risk; Uncovered time: periods when an estimated suicide risk is not available; (b) An ideal continuous-time and dynamic SRB risk prediction paradigm. In this paradigm, risks are dynamically modeled to fluctuate naturally, i.e., for every time point, the model produces an estimated SRB risk for a given prediction window following that time point, thus reducing errors associated with static approach. The continuous-time approach also provides coverage of the entire time trajectory (i.e., no “uncovered time”). t_update_: the time when new information, such as a life event, is recorded and incorporated into the risk model. Notably, if data from sources outside healthcare settings is available, t_update_ can occur independently of hospital visits, allowing for more timely updates based on the information source. Similarly, equating the timing of SRB events with the recording of diagnosis codes can introduce inaccuracies in the modeled trajectories. Further details are discussed in the Discussion section.

In this study, we aimed to develop and validate a first iteration of continuous-time, dynamic risk prediction models for SRBs. We utilize data from a large-scale electronic health records (EHR) database, encompassing more than 1.7 million patients with a wide range of demographic and clinical features, to employ a recent advance in artificial intelligence (AI): Neural Ordinary Differential Equations (Neural ODEs) ^11^. Neural ODEs are a type of deep learning model that perceives the sequential evolution of data as continuous-time differential equations, allowing for more flexible and efficient modeling of complex, dynamic systems. Specifically, our models leverage GRU-ODE^12^, an innovation that extends the original Neural ODE framework by building on Gated Recurrent Units (GRUs)^13^, a specialized type of recurrent neural network. GRU-ODE is particularly well-suited for modeling the irregularly spaced and sparse longitudinal trajectories such as those captured in her data. We further adapt GRU-ODE and develop two closely related model classes for event-based predictions (e.g., a recorded SRB), which we term “Event-GRU-ODE” and “Event-GRU-Discretized.” In this framework, each patient’s SRB risk at each time step is modeled as a non-linear transformation of a latent, continuous, and dynamic trajectory. Conceptually, such a trajectory can be interpreted as a series of abstract vectors representing the evolution of mental states), enabling the estimation of time-varying probabilities of SRBs within any specified future time window. These models have an “interleaving” design – i.e., they consist of two components— one that models a base latent trajectory of risk, and another that updates the trajectory based on information acquired from new observations. Both Event-GRU-ODE and Event-GRU-Discretized are capable of producing continuous-time predictions, with the only difference being that the former imposes an additional continuity assumption on the modeled latent trajectory – i.e., in the absence of new information, the change in the modeled value of the latent trajectory cannot exceed a threshold with respect to each time interval (see Methods for details). In short, these models can estimate continuous changes in risk across the continuum of future time points, even in the absence of new observations, and can be updated when new observations become available. By extending SRB risk estimates across the continuum of time, this approach effectively addresses one of the major limitations of current SRB risk assessment methods and could serve to inform more responsive interventions that are better able to address the complex nature of suicide risk.

## RESULTS

### Patient Cohort Characteristics

Data were obtained for 1,706,417 patients from the Research Patient Data Registry (RPDR)^14^, the EHR data repository of the Mass General Brigham (MGB) Healthcare system. From this patient set, 1,536,179 were randomly sampled for model development (80% of all patients for training, and 10 % for validation/hyperparameter tuning), and 170,238 patients (10%) were included as a hold-out test set. Table 1 summarizes the demographic composition of the data sets. Overall, the patients were predominantly females (i.e., 58% females in both training and test sets), and a majority self-identified as White (77.8% in the training/validation sets, and 77.6% in the test set) and were in the age range of 45-65 years old (Table 1 and Supplementary Figure 1). As shown in Supplementary Figure 2, most patients had fewer than 50 healthcare encounters within our “study time frame” (i.e., between Jan. 1, 2016 and Dec. 31, 2019).

**Table 1.** Demographic breakdown of the development (training and validation) and test sets.

| Setting | Training |  | Validation |  | Testing |  |
| --- | --- | --- | --- | --- | --- | --- |
|  | N | % | N | % | N | % |
| *Gender* |  |  |  |  |  |  |
| Female | 792,786 | 58.1 | 98,770 | 57.9 | 98,660 | 58.0 |
| Male | 572,798 | 41.9 | 71,787 | 42.1 | 71,573 | 42.0 |
| Unknown | 33 | 0.0 | 5 | 0.0 | 5 | 0.0 |
| *Age group* |  |  |  |  |  |  |
| < 25 y/o | 202,003 | 14.8 | 25,421 | 14.9 | 25,132 | 14.8 |
| 25–45 y/o | 382,994 | 28.1 | 47,543 | 27.9 | 47,704 | 28.0 |
| 45–65 y/o | 463,454 | 33.9 | 58,010 | 34.0 | 57,966 | 34.0 |
| > 65 y/o | 317,166 | 23.2 | 39,588 | 23.2 | 39,436 | 23.2 |
| *Race* |  |  |  |  |  |  |
| Asian | 63,510 | 4.7 | 8,029 | 4.7 | 7,876 | 4.6 |
| Black/African American | 86,681 | 6.3 | 10,739 | 6.3 | 10,850 | 6.4 |
| White | 1,062,099 | 77.8 | 132,638 | 77.8 | 132,141 | 77.6 |
| Other | 108,984 | 8.0 | 13,600 | 8.0 | 13,827 | 8.1 |
| Unknown | 44,343 | 3.2 | 5,556 | 3.3 | 5,544 | 3.3 |
| *Marital status* |  |  |  |  |  |  |
| Single | 493,167 | 36.1 | 61,958 | 36.3 | 61,264 | 36.0 |
| Married | 673,403 | 49.3 | 83,912 | 49.2 | 84,310 | 49.5 |
| Partner | 8,135 | 0.6 | 993 | 0.6 | 1,024 | 0.6 |
| Divorced | 73,209 | 5.4 | 9,163 | 5.4 | 9,226 | 5.4 |
| Separated | 12,366 | 0.9 | 1,530 | 0.9 | 1,504 | 0.9 |
| Widowed | 58,504 | 4.3 | 7,188 | 4.2 | 7,189 | 4.2 |
| Other/unknown | 46,833 | 3.4 | 5,818 | 3.4 | 5,721 | 3.4 |
| *Veteran status* |  |  |  |  |  |  |
| Yes | 66,325 | 4.9 | 8,280 | 4.9 | 8,360 | 4.9 |
| No | 1,081,395 | 79.2 | 135,128 | 79.2 | 134,750 | 79.2 |
| Unknown | 217,897 | 16.0 | 27,154 | 15.9 | 27,128 | 15.9 |
| *Public payor* |  |  |  |  |  |  |
| Yes | 627,056 | 45.9 | 78,060 | 45.8 | 78,392 | 46.0 |
| No | 738,561 | 54.1 | 92,502 | 54.2 | 91,846 | 54.0 |
| *Income level* |  |  |  |  |  |  |
| < \$40k | 96,897 | 7.1 | 12,094 | 7.1 | 12,027 | 7.1 |
| \$40k–\$70k | 572,996 | 42.0 | 71,915 | 42.2 | 71,381 | 41.9 |
| \$70k–\$100k | 476,069 | 34.9 | 59,451 | 34.9 | 59,499 | 35.0 |
| > \$100k | 219,655 | 16.1 | 27,102 | 15.9 | 27,331 | 16.1 |

Our models continuously generate predictions on a daily basis for each patient, with each prediction varying in its temporal distance from both the patient’s first recorded observation and the last observation before the prediction was made. We illustrate the distribution of the number of months between the prediction timepoint and (1) the first observation (representing the total length of information available for the prediction in question) and (2) the last available observation in Supplementary Figure 3. Most predictions were made with less than 2 years of data from the first observation and less than 5 months from the last observation. See below for the effect of length of observed patient history on model performance.

### Main Model Performance Metrics

Table 2 shows the prediction performance of the Event-GRU-ODE and Event-GRU-Discretized models. In general, both models achieved excellent discrimination (area under the receiver operator curve, AUROC > 0.9) across different prediction windows (ROC curves are shown in Supplementary Figure 4). All model metrics were reported at a fixed 95% specificity. The highest AUROCs were recorded with a 1-month prediction window (Event-GRU-ODE=0.940, Event-GRU-Discretized=0.942), while the highest area under the precision-recall curve (AUPRC) and positive predictive value (PPV) were observed using a 1.5-year prediction window, for both models (AUPRC=0.089 and PPV=0.016 for Event-GRU-ODE; AUPRC=0.089 and PPV=0.017 for Event-GRU-Discretized). All AUPRCs and PPVs were less than 0.09 and 0.02, respectively, due to the low SRB prevalence in the data (0.01% - 0.12%). Although we observed the lowest PPV with the shortest prediction window, those classified as high risk were 15 times (RR=15.16) more likely to have an SRB than those classified as low risk. While both models had nearly identical AUROC, AUPRC and PPV, the Event-GRU-Discretized model achieved slightly better results in sensitivity and relative risk (at 95% specificity) than Event-GRU-ODE, with longer prediction windows (e.g., 1 year or 1.5 years).

**Table 2.** Model performance comparison between Event-GRU-ODE (i.e., “ODE” in the table) and Event-GRU-Discretized (i.e., “Discretized” in the table), for the five prediction windows (Pred. win.). Performance metrics are AUROC, AUPRC, positive predictive value (PPV), sensitivity and relative risk (RR = PPV/prevalence), reported at 95% specificity.

| Model | Prediction Window | AUROC | AUPRC | PPV | Sensitivity | RR | Prevalence |
| --- | --- | --- | --- | --- | --- | --- | --- |
| ODE | 1 month | 0.940 | 0.023 | 0.002 | 0.759 | 15.16 | 0.01% |
| Discretized | 1 month | 0.942 | 0.024 | 0.002 | 0.759 | 15.16 | 0.01% |
| ODE | 3 months | 0.935 | 0.046 | 0.005 | 0.732 | 14.58 | 0.03% |
| Discretized | 3 months | 0.938 | 0.044 | 0.005 | 0.735 | 14.64 | 0.03% |
| ODE | 6 months | 0.933 | 0.062 | 0.008 | 0.701 | 13.91 | 0.06% |
| Discretized | 6 months | 0.936 | 0.061 | 0.008 | 0.718 | 14.25 | 0.06% |
| ODE | 1 year | 0.927 | 0.080 | 0.014 | 0.675 | 13.32 | 0.11% |
| Discretized | 1 year | 0.930 | 0.080 | 0.014 | 0.694 | 13.70 | 0.11% |
| ODE | 1.5 years | 0.925 | 0.089 | 0.016 | 0.668 | 13.16 | 0.12% |
| Discretized | 1.5 years | 0.929 | 0.089 | 0.017 | 0.688 | 13.55 | 0.12% |

### Effect of Time Length of Observed Patient History on Model Performance

Figure 3 plots model metrics for the Event-GRU-ODE model as a function of the time length of observed patient history, smoothed by the locally weighted scatterplot smoothing (LOWESS) method and bootstrapped 95% confidence intervals. In general, performance tended to be better with longer observed patient history trajectories (and, therefore, more accumulated information), except for the AUPRC curve using the 1-month prediction window. We observe wider confidence intervals around the LOWESS fits towards the end of the follow-up period and with longer prediction windows. This widening may be attributed to a combination of factors: patients moving in and out of the hospital system, resulting in not all being followed for the entire study period; and the exclusion of time steps from the model training when their prediction windows extend beyond the study period. Both of these factors contribute to a reduction in the amount of data available for training during the later time steps, relative to the initial records of each patient. The aggregated prediction plots for the discretized model are shown in Supplementary Figure 5.

### Subgroup Analysis by Clinical Settings and Demographic Factors of Interest

Figure 4 presents model metrics for Event-GRU-ODE stratified by three different clinical settings: (1) “General,” including all available patients and visits; (2) “Psych ED,” for which predictions were made at the time of an emergency department visits involving psychiatric evaluation/consultation; and (3) “Psych Inpatient,” for which predictions were made at time of a psychiatric inpatient admission. AUPRC and PPV were significantly higher for the “Psych ED” and “Psych Inpatient” cohort compared to the “General” cohort, presumably due to the increased prevalence of SRB events among the two sub-cohorts the across different prediction windows (2.43% - 11.38% for “Psych ED”, and 0.78% - 7.24% for “Psych Inpatient”). The highest PPV (42.2%) was achieved for the Psych ED setting, with a prediction window of 1.5 years.

Event-GRU-ODE performance metrics stratified by important patient demographic characteristics (i.e., gender, self-reported race, age, income, and public payor for healthcare) are shown in Supplementary Figures 6-10. In general, AUROCs demonstrated modest variation across different demographic groups but were good (AUROC > 0.8) in all settings except for longer prediction windows among those with age < 20. There was greater variation in AUPRCs which were highest among individuals whose self-reported race was Black (for prediction windows of 6 months or more), and also tended to be higher among individuals aged 20-60 years and those gender was male. The discrete model version demonstrated similar characteristics in both subgroup analyses (data not shown).

### Effect of Training Data Size on Model Performance

The size of available training data can be a limiting factor in settings with constrained resources. Theoretically, if the continuity assumption underlying the Event-GRU-ODE model holds true, this assumption should benefit model performance when training data is limited. To examine the overall effect of training data size on performance across both model versions, as well as the impact of the continuity assumption (demonstrated by contrasting the ODE and discretized models), we present in Table 3 the results (in AUROC and AUPRC) from training both models with various data sample sizes (i.e., 1/8, 3/8, 5/8, and 8/8 or 100% of the training set). Additionally, we plot the comparison of prediction performance (in AUROC, AUPRC, and PPV at 95% specificity) between the ODE and discretized models for the 3- month prediction in Figure 2. We choose the 3-month (i.e., 90-days) prediction window due to the clinical utility of shorter-term prediction intervals. In general, model performance was only modestly reduced when training data was 3/8 (37.5%) of the full training dataset. We observe a more significant decline in model performance when the training data size is reduced from 3/8 to 1/8 of all available training data. However, discriminative metrics were still robust (AUROC > 0.84 for both model classes) under the 1/8 training data setting. Of note, Event-GRU-ODE achieved higher AUPRC and PPV (a model metric which have been shown to be more informative than AUROC when base prevalence is low^15,16^) compared to Event-GRU-Discretized with smaller training data sizes, i.e., with only 3/8 or 1/8 of training data. Performance plots for the other prediction windows are shown in Supplementary Figures 11-14.

**Figure 2.**
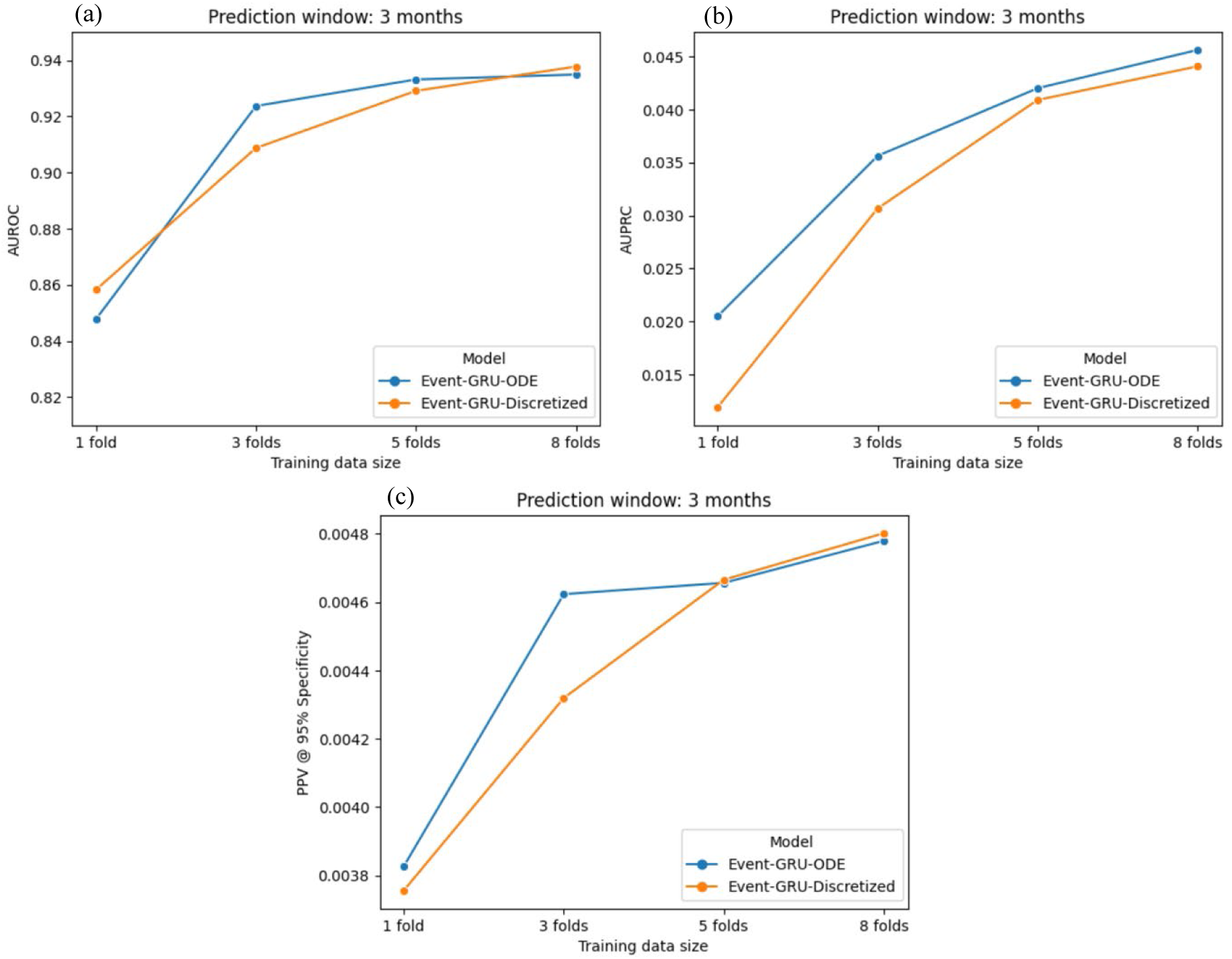
Model performance (in (a) AUROC; (b) AUPRC; and (c) PPV at 95% specificity) comparison between the two models trained with different proportions of the training data (1 fold, 3 folds, 5 folds and 8 folds, i.e., 1/8, 3/8, 5/8, and all of the training set), using a 3-month prediction window.

**Table 3.** AUROC and AUPRC scores of models trained using different proportions of the data set (1 fold, 3 folds, 5 folds and 8 folds, which are 1/8, 3/8, 5/8 and 100% of the training set, respectively).

| Model | Metric | Prediction Window | Fraction of data used for training |  |  |  |
| --- | --- | --- | --- | --- | --- | --- |
|  |  |  | 1 fold | 3 folds | 5 folds | 8 folds (all) |
| ODE | AUROC | 1 month | 0.862 | 0.928 | 0.937 | 0.94 |
| ODE | AUPRC | 1 month | 0.009 | 0.017 | 0.024 | 0.023 |
| ODE | AUROC | 3 months | 0.848 | 0.924 | 0.933 | 0.935 |
| ODE | AUPRC | 3 months | 0.02 | 0.036 | 0.042 | 0.046 |
| ODE | AUROC | 6 months | 0.841 | 0.923 | 0.932 | 0.933 |
| ODE | AUPRC | 6 months | 0.034 | 0.049 | 0.056 | 0.062 |
| ODE | AUROC | 1 year | 0.824 | 0.917 | 0.926 | 0.927 |
| ODE | AUPRC | 1 year | 0.044 | 0.063 | 0.071 | 0.08 |
| ODE | AUROC | 1.5 years | 0.822 | 0.916 | 0.925 | 0.925 |
| ODE | AUPRC | 1.5 years | 0.047 | 0.07 | 0.078 | 0.089 |
| Discretized | AUROC | 1 month | 0.872 | 0.917 | 0.935 | 0.942 |
| Discretized | AUPRC | 1 month | 0.006 | 0.018 | 0.023 | 0.024 |
| Discretized | AUROC | 3 months | 0.858 | 0.908 | 0.929 | 0.938 |
| Discretized | AUPRC | 3 months | 0.012 | 0.031 | 0.041 | 0.044 |
| Discretized | AUROC | 6 months | 0.848 | 0.907 | 0.928 | 0.936 |
| Discretized | AUPRC | 6 months | 0.018 | 0.039 | 0.056 | 0.061 |
| Discretized | AUROC | 1 year | 0.835 | 0.897 | 0.922 | 0.93 |
| Discretized | AUPRC | 1 year | 0.025 | 0.046 | 0.073 | 0.08 |
| Discretized | AUROC | 1.5 years | 0.832 | 0.896 | 0.921 | 0.929 |
| Discretized | AUPRC | 1.5 years | 0.028 | 0.05 | 0.081 | 0.089 |

## DISCUSSION

In this study, we present a continuous-time, dynamic modeling approach for predicting SRB risks. Unlike current assessment methods, this approach accommodates the time-varying nature of SRB risks and, by generating risk estimates across the entire time trajectory, it mitigates gaps in risk score availability. We utilized data from a large-scale EHR database containing data from multiple medical centers and hospitals, including features extracted from unstructured clinical notes, to develop and validate two model classes (Event-GRU-ODE and Event-GRU-Discretized) based on an advanced neural network architecture - Neural ODEs. Both model classes achieved high discrimination performance across different clinical settings. For example, in the general setting, AUROCs were between 0.93-0.94, and relative risks (the modeled risk divided by baseline prevalence during the designated prediction window) ranged from 13.2 and 15.2 across different prediction windows at 95% specificity. Such performance is better or comparable to those observed in previous analyses based on conventional static risk estimates by our group and others^4,6–10,17–19^.

With respect to analyses by clinical setting, the results largely tracked the base rate of SRBs. As expected, AUROCs were generally higher and PPVs lower for the “General” (all patients) cohort than the inpatient or ED sub-cohorts where base rates are higher. Stratified analyses by demographics found good discrimination across groups (AUROCs > 0.80) while AUPRCs were more variable (again, consistent with varying base rates of the outcome). For prediction windows of 6 months or longer, AUPRCs tended to be highest among those who identified as Black and lowest among those who identified as Asian.

While the two model classes achieved similar performance across various metrics when all training data were used, Event-GRU-ODE attained higher AUPRC and PPVs than its discretized counterpart for the majority of configurations in the settings where the size of training data is limited. This implies that the continuity prior assumption^12^ (see Methods) may be beneficial in such settings.

While these proof-of-concept analyses focused solely on EHR data for model development, a key advantage of our model classes lies in their flexible architecture, including their natural incorporation of a time dimension. This flexibility facilitates the easy integration of additional data modalities, such as genetic data or other biomarkers, and enables updates to the modeled future risk trajectory based on new incoming information at any point in time. This is crucial because, although a risk trajectory generated at a certain point can theoretically cover all future times, it may diverge from reality if it extends too far into the future without incorporating updated information. Sources of new information can include future healthcare visits, data from digital sensors, or outputs from mobile devices, which may themselves be structured as time series. Crucially, updating the modeled risk trajectory with incoming information at a specific time step requires only the modeled latent state vector from the previous time step, the relevant new information content, and the model parameters. This approach eliminates the need for access to the original data utilized to generate the risk trajectory up to the current time point, which may contain protected health information, thereby mitigating privacy concerns when updating predictions outside healthcare facilities.

Our results should be interpreted with several limitations in mind. Firstly, in this initial iteration of continuous-time SRB prediction models, our models rely solely on EHR data, with limited use of information contained within clinical notes (i.e., only utilizing the presence of specific extracted terms). A natural next step would be to refine the precision of event timing (including SRBs), potentially through the application of natural language processing techniques (e.g., large language models^20–22)^ in combination with the information contained in clinical notes. Moreover, integrating sources of information beyond the healthcare system could further ensure timely updates of the modeled trajectory and potentially enhance model performance. Secondly, this initial study on continuous-time SRB prediction models has not yet explored their generalizability in depth. Lastly, model interpretation in time series modeling is an evolving field. While applying standard interpretation techniques, such as SHAP (Shapley Additive Explanations)^23^, is technically possible, it often leads to complex and non-straightforward interpretations. Due to this complexity, we didn’t include an analysis of feature importance.

Effective suicide prevention strategies necessitate a collaborative approach between the healthcare system and the broader community, including schools, workplaces, and families. The long-term goal of developing risk prediction tools that can adapt to dynamic changes in SRB risk, integrate new information continuously, and offer timely assessments, is to enhance the design of suicide prevention strategies. For example, data from community settings could be integrated into real-time risk assessments for more timely delivery of support or interventions. Alternatively, healthcare visits might be strategically scheduled during periods of predicted spikes in risk, facilitating better planning around times of the year or following specific life events that may elevate risk.

In conclusion, leveraging neural ODE—a recent advance in deep learning for time-series analysis—we have developed continuous-time and dynamic risk prediction models to assess the risk of SRB using EHR data. We validated their effectiveness across various prediction windows, with robust performance across diverse demographics and clinical settings. Our results confirm the feasibility of continuous-time and dynamic prediction for SRB and highlight the value of continued innovation in this area to enable more accurate and comprehensive SRB risk stratification methods.

## METHODS

### Data source and study population

The data used for this study were derived from RPDR^14^ , the research EHR data registry of the MGB healthcare system located in Massachusetts, USA. The RPDR is a centralized data registry that collects and compiles clinical information from multiple EHR systems within MGB, which includes more than 7 million patients with over 3 billion records seen across more than 8 hospitals, including two major teaching hospitals^14^. In this study, we used data between the period of Jan. 1, 2016 and Dec. 31, 2019 (the “study time frame”) and included all patients who had at least 3 visits and a minimum of 30 days of medical record, and were older than age 10 at the time of their first medical record entry within this time period. We chose the start date to minimize the impact of data heterogeneity caused by different recording practices due to the hospital system’s transition from ICD (International Classification of Disease)-9 to ICD-10.

### Suicide-related behavior definition

For the purpose of this study, we adopt a list of ICD codes, previously developed and validated^8,24^, as the definition of SRB. These codes were shown to have to be valid for capturing intentional self-harm by extensive chart review by expert clinicians, a PPV of greater than 70%^24^. An “SRB event” is then defined by the occurrence of any of the codes in the list.

### Predictors

We include two types of predictors: (1) *covariates*, including the demographic variables (total of 7, a full list is provided in Table 1). These enter the model at the beginning of the trajectories; and (2) *observations*, which are recorded for each hospital encounter during the patient trajectory, including: a. diagnosis codes, which were mapped from ICD codes^25,26^ to PheWAS codes^27^ which have been shown to capture clinically meaningful concepts^28^ (total 1,871 features); b. medications, which were mapped to RXNORM codes^29^ (total 1,712 features); and c. a set of previously defined features derived using natural language processing (NLP)^30,31^, which we named as “NLP CUIs” (i.e., concept unique identifiers^32^), indicating the presence of mental health-related concepts as defined in the Unified Medical Language System (UMLS^32^) in the clinical notes (total of 2,488 features). Observations were ordered temporally and enter the model as inputs according to their recorded date relative to the date of the first entry of each patient (see Figure 5 for a schematic illustration of the data setup). Note that while we chose a time resolution of one day as the basic unit for a time step, any time resolution can theoretically be chosen given that data are available (the smaller the time step, the closer the approximation to continuity).

**Figure 3.**
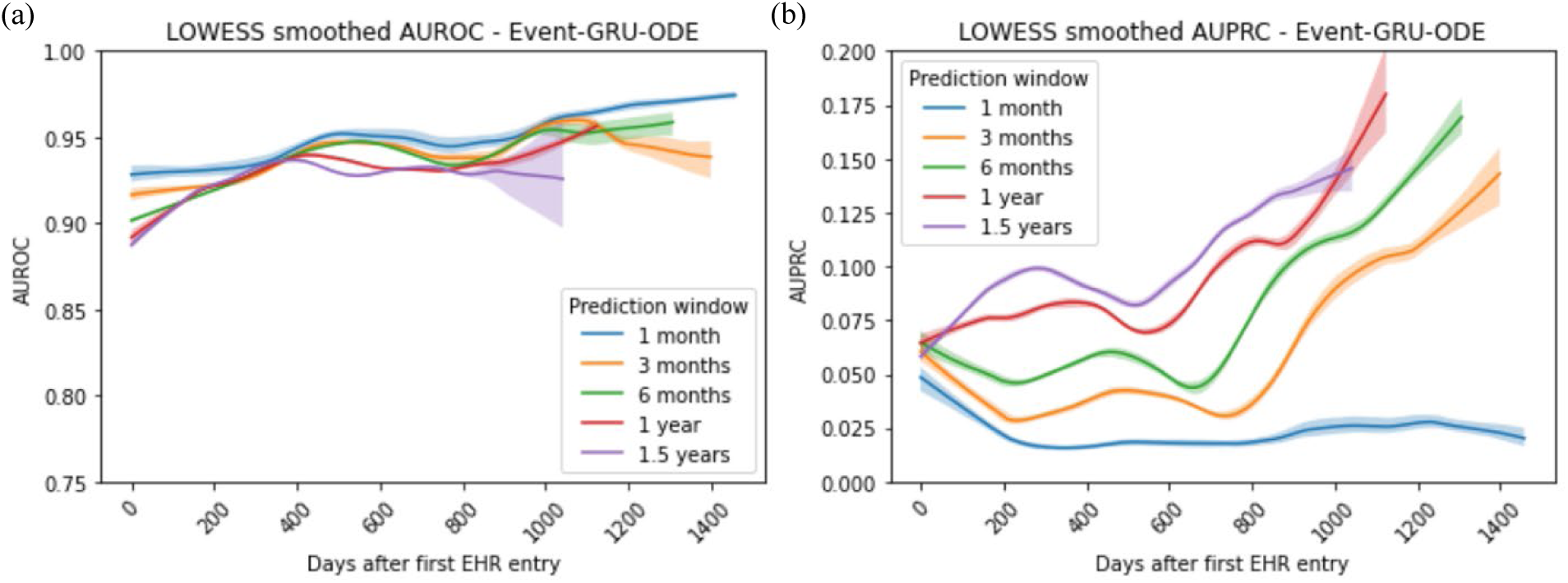
Aggregated prediction performance (in (a) AUROC and (b) AUPRC) by the length of observed history (smoothed by LOWESS) since the beginning of every patient trajectory, for Event-GRU-ODE.

**Figure 4.**
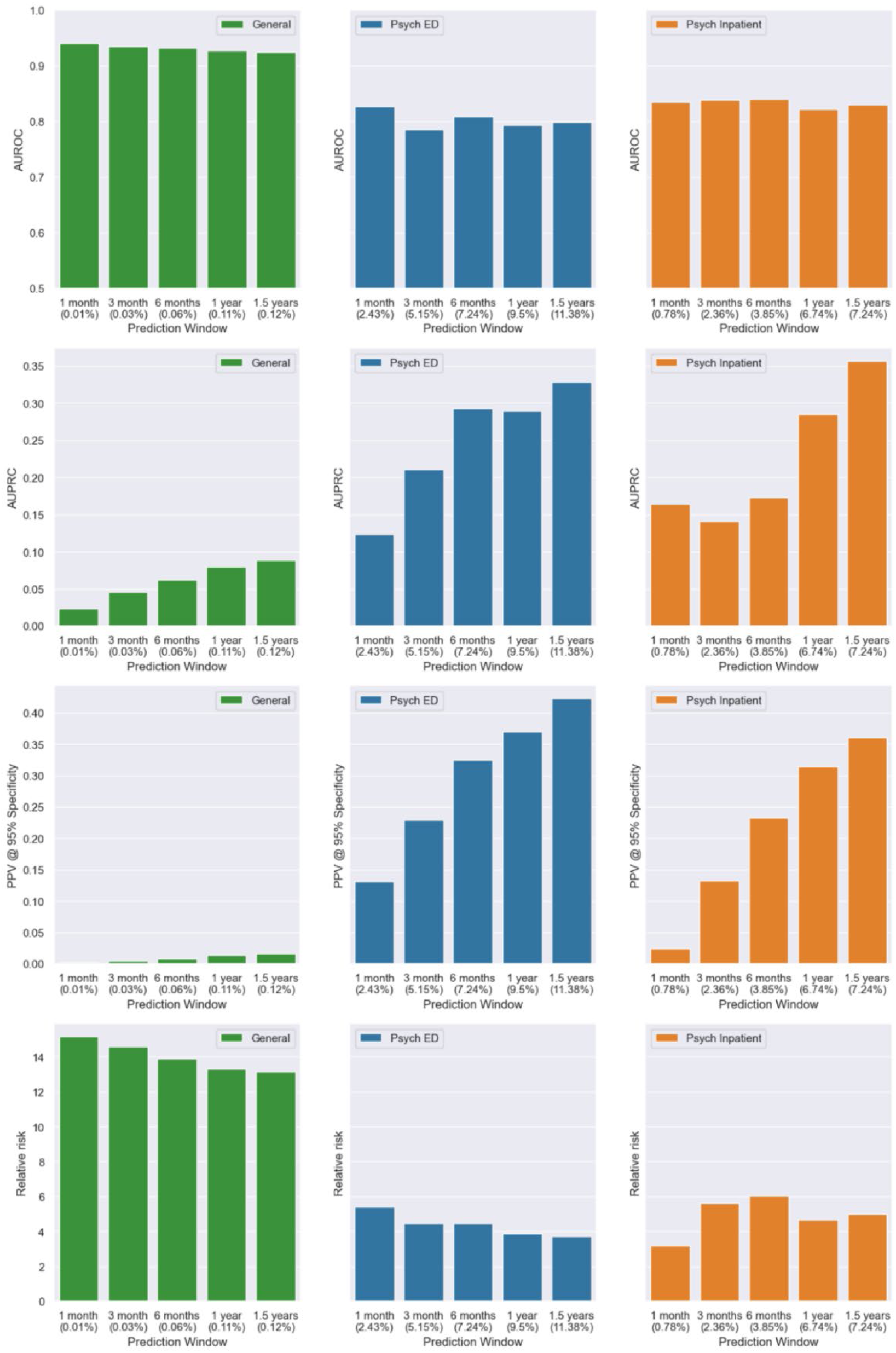
Model performance by clinical settings, namely “General” (general setting, green), “Psych ED” (psychiatric emergency department, blue) and “Psych Inpatient” (psychiatric inpatients, orange), for Event-GRU-ODE. The percentages in the parentheses along the x-axis indicate the prevalence of SRB in each setting.

**Figure 5.**
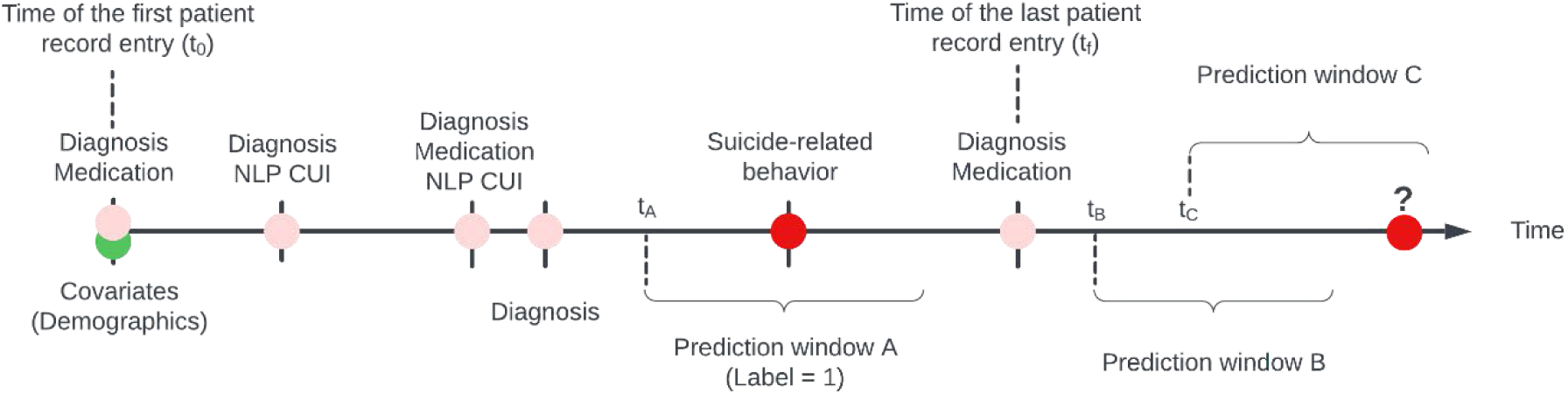
Schematic diagram of the data setup for one hypothetical patient. Static covariates (demographic observations), which may include any combination of diagnoses, medications, and NLP CUIs, are entered into the model at time t0 (i.e., the time of the first patient entry). Subsequent observations are ordered sequentially over time, with the exact time steps (days after the first record) preserved. Labels are created for each time step, considering the presence of a SRB record. For any time step t_A_ within the interval where data is available, its associated prediction Window A, starting from t_A_+1, is assigned a label value of 1 if SRB is recorded within that period. Note that t_A_ is not bounded to the dates of where there are recorded entries. With the trained model, predictions can be made for any future time step (t_B_, t_C_, …) occurring after the last recorded entry at t_f_ (note that labels are not available for future times and the probability of their occurrences are the targets for predictions (red circle with question mark)).

Continuous-valued features (e.g., age range at the time follow-up started) were discretized by binning, and all features were converted to one-hot encoding before entering the models.

### Prediction task definition

Based on the SRB definition, we formulate the suicide risk prediction task as follows:

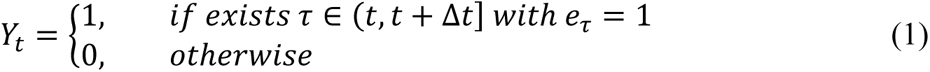

where Δ*t* is the length of the “prediction window” and *e_τ_* is 1 if an event (i.e., suicide-related behavior) was recorded at time *τ*. This means *Y_t_* (i.e., the prediction label) is set to 1 if an event happens within the next Δ*t* time (i.e., a “prediction window”). In this study, we look at prediction windows of 1 month, 3 months, 6 months, 1 year, and 1.5 years.

Prediction labels are created for each patient, for each day, from the date of the first visit up to the date of the last visit, minus the length of the prediction window to ensure the full length of prediction window is observed. Of note, to compensate for the fact that events recorded toward the end of patient history would contribute fewer labels under this scheme, a “buffering time” of 30 days is appended to the date of the last visit, during which the label of the last visit will be carried over up to the 30^th^ day after the last visit.

### Event-GRU-ODE model

We model a patient trajectory as a *D*-dimensional stochastic process ***Y***(*t*) whose dynamics is driven by a stochastic differential equation (SDE):

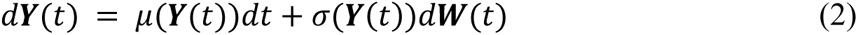

where d***W***(t) is a Wiener process. The EHR records in a patient trajectory typically consist of sparse and irregularly observed longitudinal data, which makes modeling the SDE directly very difficult.

We follow the idea from statistical mechanics, that instead of modeling SDE directly, one models the time evolution of the probability density function of the observables. Therefore, we propose to use a latent variable process ***h***(*t*) that can be mapped to the mean and covariance of the target variables of interest, i.e., the SRB labels ***Y***_t_ (see “Prediction task definition”). In detail, we model the probabilities of a SRB event happening between time period *t* and *t* + Δ*t*, in the form of a Bernoulli distribution. This means that the latent process ***h***(*t*), varying in time, is mapped to the parameter *p*(*t*) of the Bernoulli distribution, which determines its mean and variance varying in time. The dynamics of the underlying latent process are governed by ordinary differential equations (ODEs), whose parameters are learned from data.

Specifically, we model the mapping from the observed variables to the hidden variable with “Event-GRU-ODE” as described below, and use a feed-forward neural network to map the hidden variable ***h***(*t*) to the output probabilities (i.e., the parameter *p* of Bernoulli distribution of the random variable). This SDE formulation reflects the evolving uncertainty of the patient’s future condition regarding SRB.

#### Model components

To model the latent variable ***h***(*t*), used for predicting the probabilities of SRB events, and incorporate incoming health-related observations, such as diagnosis and measurements, we propose a two-mode system, consisting of: (1) *Continuous time propagation* of the latent variable ***h***(*t*); and (2) *Discrete updates* that happen each time a new observation *x_t_* is received at time *t*. Each time there is an observation that triggers the discrete update the system switches to a new state of the ODE (which still has the same parameters and dynamics as before).

#### Continuous time propagation

For the *continuous time propagation*, we employ recently proposed methods in the field of Neural ODEs, specifically GRU-ODE^33^. This means when there are no observations, the latent variable follows an ODE whose trajectory is controlled by the GRU-ODE:

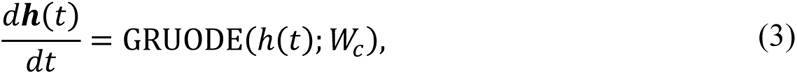

where *W_c_* are the weights of the neural networks of GRU-ODE (learned end-to-end with other components of the model).

To derive GRU-ODE we start with the standard classic GRU^13^, given by its equations for update gate *z*, reset gate r and candidate hidden vector 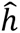:

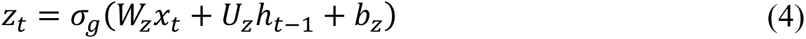

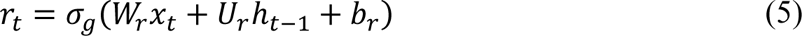

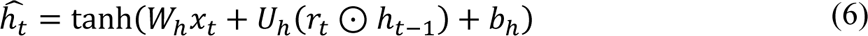

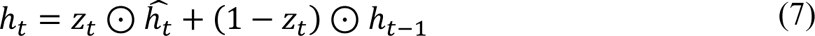

The key idea is to observe that the last equation, which combines the candidate hidden vector 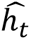 and the previous hidden vector ***h***_*t*−1_, can be rewritten as a difference equation for ***h***_*t*_:

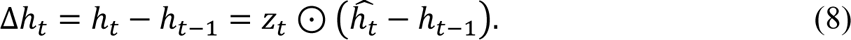

Taken to the limit of Δ*t* → 0 the difference equation gives an ODE with negative feedback on the current hidden state ***h***(*t*):

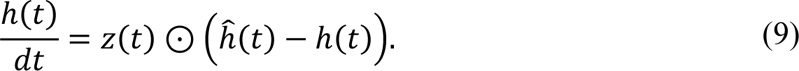

GRU-ODE has several useful properties, such as stability due to negative feedback and Lipschitz continuity with respect to time with constant *K* = 2, which helps make the model robust to numerical errors as well as effective even with smaller data sample size^33^.

#### Initial hidden state

The initial hidden state, ***h***(0), is computed from the static covariates of the patient using a multilayer feed-forward neural network.

#### Discrete update

The continuous-time GRU-ODE component propagates the latent vector up to the observation time (*t*) where the model then updates the hidden state ***h***(*t*) to a new value ***h***′ by using a discrete network, which is a standard GRU^13^:

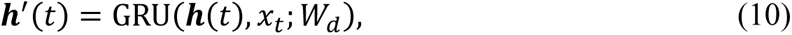

where *x_t_* is the observation at time *t* and *W_d_* are the weights of the update GRU network and a multilayer feed-forward network that preprocesses *x_t_*. After this update the system switches back to continuous-time propagation (GRU-ODE) starting from the state ***h***^′^(*t*).

Finally, we use an output network *f*(*h*(*t*); *W_o_*) that maps the hidden state *h*(*t*) to the SRB event probabilities *p*(*t*) for the time *t*. Using *cross-entropy loss* on the outputs of the network and feeding in observations *x_t_* we can train Event-GRU-ODE in fully *end-to-end manner*.

### Event-GRU-Discretized model

The Event-GRU-ODE model makes the assumption that between two observations (e.g., two healthcare encounters) the latent vector ***h*** is propagating by following an ODE, forcing the trajectory to be continuous in time.

We can relax this continuity assumption by allowing the time propagation to be updated by the discrete GRU. This propagation is still carried out in iterative fashion using time step size as in Event-GRU-ODE. Relaxing the continuity assumption could allow the model to be more flexible and might affect performance. We name this model “Event-GRU-Discretized.”

### Model training and evaluation

We implemented and evaluated two models, namely “Event-GRU-ODE” and “Event-GRU-Discretized”, for the continuous time suicide risk prediction task, with 5 different prediction windows (i.e., 1 month, 3 months, 6 months, 1 year and 1.5 years). We randomly split the data into 10 “folds,” each contains data from 10% of all patients. Among them, 8 folds (80%) were used as a training set, one fold (10%) as a hold-out validation set (10%) for tuning all the model hyperparameters, and one fold (10%) as a hold-out test set for model evaluation (10%). During training, models were optimized using the Adam optimizer^34^. More detailed training procedures, including hyperparameter settings, are provided in Supplementary Note A.

For model performance evaluation, we report area under the receiver operator characteristic curve (AUROC), area under Precision-Recall curve (AUPRC), as well as positive predictive value (PPV) and sensitivity with specificity set to 0.95.

In order to evaluate how well Event-GRU-ODE makes its predictions as a function of the length of observed patient history trajectories, we aggregated all the model predictions made on each day after the first EHR entry of each patient, measured model performance (in AUROC and AUPRC) along the time trajectory, and plotted the model metrics over time for the study time frame. To increase the signal-to-noise ratio, plots were smoothed by the LOWESS method with its smoothing window (i.e., the fraction parameter) set to be 0.3. We also performed bootstrap resampling to add the estimated 95% confidence intervals around each LOWESS fit on the plot.

In addition, we assessed potential differences in model performance across various patient subgroups. First, we defined three patient groups of interest based on clinical settings to stratify prediction results and evaluate model performance in each patient population: (1) “General” representing the general outpatient cohort, which encompasses the full study population and their complete trajectories within the study period; (2) “Psych ED” for predictions during emergency department visits involving psychiatric evaluation/consultation within the study period; (3) “Psych Inpatient” for predictions made during psychiatric inpatient admissions within the study period. Furthermore, we conducted stratified analyses on key demographic groups (gender, race, age group, income level by ZIP code, whether the patient has a public insurance payor) to examine potential heterogeneity in model performance across different subgroups.

Lastly, to further compare the two models and their capabilities to learn from fewer samples, we trained each model using all the training data as well as randomly sampling 5/8, 3/8 and 1/8 (i.e., 5, 3, and one fold, respectively) of the training data.

## DATA AVAILABILITY

Protected Health Information restrictions apply to the availability of the clinical data here, which were used under IRB approval for use only in the current study. As a result, this dataset is not publicly available.

## ACKNOWLEDGEMENTS

Drs. Smoller, Sheu, and Mr. Lee were supported in part by NIMH R01 MH117599.

## AUTHOR CONTRIBUTIONS

Y.-H.S. conceived of the study; Y.-H.S. and J.W.S. acquired the data for the study; J.S. developed the models utilized in the study; Y.-H.S., B.W., and H.L. curated data and performed analyses of the study; J.W.S., supervised analyses of the study; J.W.S. provided computational resources for the study; Y.-H.S., J.S., and B.W. wrote the first draft; and all authors revised the manuscript and contributed to interpretation of the analyses of the study.

