## Supplementary materials for "Continuous-Time and Dynamic Suicide Attempt Risk Prediction with Neural Ordinary Differential Equations"

Supplementary On-line Materials


| | Page |
|---|---|
| **Supplementary Table 1** | **3** |
| **Supplementary Figure 1** | **5** |
| **Supplementary Figure 2** | **6** |
| **Supplementary Figure 3** | **7** |
| **Supplementary Figure 4** | **8** |
| **Supplementary Figure 5** | **9** |
| **Supplementary Figure 6** | **10** |
| **Supplementary Figure 7** | **11** |
| **Supplementary Figure 8** | **12** |
| **Supplementary Figure 9** | **13** |
| **Supplementary Figure 10** | **14** |
| **Supplementary Figure 11** | **15** |
| **Supplementary Figure 12** | **16** |
| **Supplementary Figure 13** | **17** |
| **Supplementary Figure 14** | **18** |
| **Supplementary Note A** | **20** |

### SUPPLEMENTARY TABLE

**Supplementary Table 1 |** SRB prevalence by stratified patient groups.

**Supplementary Table 1 |** SRB prevalence by stratified patient groups.

| **Subgroup** | **Prevalence (%)** | | | | |
|---|---|---|---|---|---|
| | 1 month | 3 months | 6 months | 1 year | 1.5 years |
| *Clinical setting* | | | | | |
| General | 0.013 | 0.033 | 0.059 | 0.105 | 0.124 |
| Psych ED | 2.438 | 5.152 | 7.243 | 9.505 | 11.377 |
| Psych Inpatient | 0.782 | 2.359 | 3.851 | 6.735 | 7.241 |
| *Gender* | | | | | |
| Female | 0.011 | 0.029 | 0.051 | 0.090 | 0.106 |
| Male | 0.016 | 0.039 | 0.070 | 0.128 | 0.153 |
| *Age group* | | | | | |
| < 20 y/o | 0.039 | 0.097 | 0.170 | 0.315 | 0.380 |
| 20–60 y/o | 0.016 | 0.040 | 0.072 | 0.131 | 0.156 |
| > 60 y/o | 0.004 | 0.010 | 0.018 | 0.029 | 0.032 |
| *Race* | | | | | |
| Asian | 0.009 | 0.022 | 0.040 | 0.067 | 0.079 |
| Black/African American | 0.019 | 0.047 | 0.081 | 0.143 | 0.165 |
| White | 0.013 | 0.032 | 0.058 | 0.104 | 0.123 |
| Other | 0.013 | 0.031 | 0.057 | 0.101 | 0.121 |
| Unknown | 0.013 | 0.037 | 0.071 | 0.123 | 0.149 |
| *Public payer* | | | | | |
| Yes | 0.021 | 0.052 | 0.095 | 0.168 | 0.198 |
| No | 0.006 | 0.013 | 0.023 | 0.040 | 0.047 |
| *Income level* | | | | | |
| < $40k | 0.021 | 0.051 | 0.095 | 0.187 | 0.229 |
| $40k–$70k | 0.016 | 0.042 | 0.076 | 0.136 | 0.160 |
| $70k–$100k | 0.010 | 0.025 | 0.044 | 0.077 | 0.091 |
| > $100k | 0.009 | 0.021 | 0.038 | 0.064 | 0.076 |

### SUPPLEMENTARY FIGURES

**Supplementary Figure 1 |** Distribution of patient age (at the start of their patient trajectories).

**Supplementary Figure 2 |** (a) Distribution of the length of the patient trajectories (in days). (b) Distribution of number of observations (in each patient trajectory).

**Supplementary Figure 3 |** (a) Distribution of number of months since the first observation. (b) Distribution of number of months since each patient's previous observation in the data set.

**Supplementary Figure 4 |** Receiver Operating Characteristic (ROC) curve comparison between Event-GRU-ODE and Event-GRU-Discretized, for each prediction window ((a): 1 month; (b) 3 months; (c) 6 months; (d) 1 year; (e) 1.5 years). The plots show almost identical curves obtained from the two models.

**Supplementary Figure 5 |** Aggregated prediction performance ((a) AUROC; (b) AUPRC) over time (smoothed by LOWESS) since the beginning of every patient trajectory, for Event-GRU-Discretized.

**Supplementary Figure 6 |** Model performance ((a) AUROC and (b) AUPRC) stratified by gender for Event-GRU-ODE.

**Supplementary Figure 7 |** Model performance ((a) AUROC and (b) AUPRC) stratified by self-reported race for Event-GRU-ODE.

**Supplementary Figure 8 |** Model performance ((a) AUROC and (b) AUPRC) stratified by age groups for Event-GRU-ODE.

**Supplementary Figure 9 |** Model performance ((a) AUROC; (b) AUPRC) stratified by income level by ZIP codes for Event-GRU-ODE.

**Supplementary Figure 10 |** Model performance ((a) AUROC and (b) AUPRC) stratified by whether patients have a public payor for Event-GRU-ODE.

**Supplementary Figure 11 |** Model performance comparison ((a) AUROC; (b) AUPRC; (c) PPV at 95% specificity) between the two models trained with different proportions of the data set (1 fold, 3 folds, 5 folds and 8 folds), using a 1-month prediction window.

**Supplementary Figure 12 |** Model performance comparison ((a) AUROC; (b) AUPRC; (c) PPV at 95% specificity) between the two models trained with different proportions of the data set (1 fold, 3 folds, 5 folds and 8 folds), using a 6-month prediction window.

**Supplementary Figure 13 |** Model performance comparison ((a) AUROC; (b) AUPRC; (c) PPV at 95% specificity) between the two models trained with different proportions of the data set (1 fold, 3 folds, 5 folds and 8 folds), using a 1-year prediction window.

**Supplementary Figure 14 |** Model performance comparison ((a) AUROC; (b) AUPRC; (c) PPV at 95% specificity) between the two models trained with different proportions of the data set (1 fold, 3 folds, 5 folds and 8 folds), using a 1.5-year prediction window.

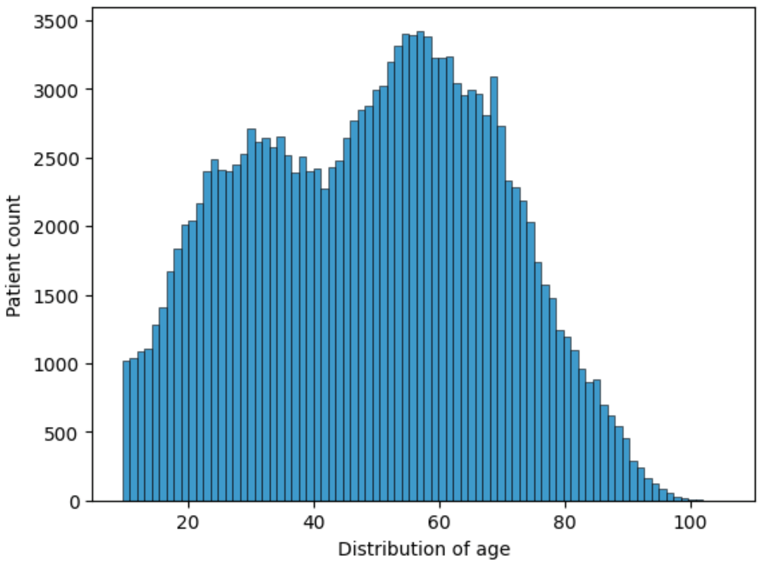


**Supplementary Figure 1 |** Distribution of patient age (at the start of their patient trajectories).

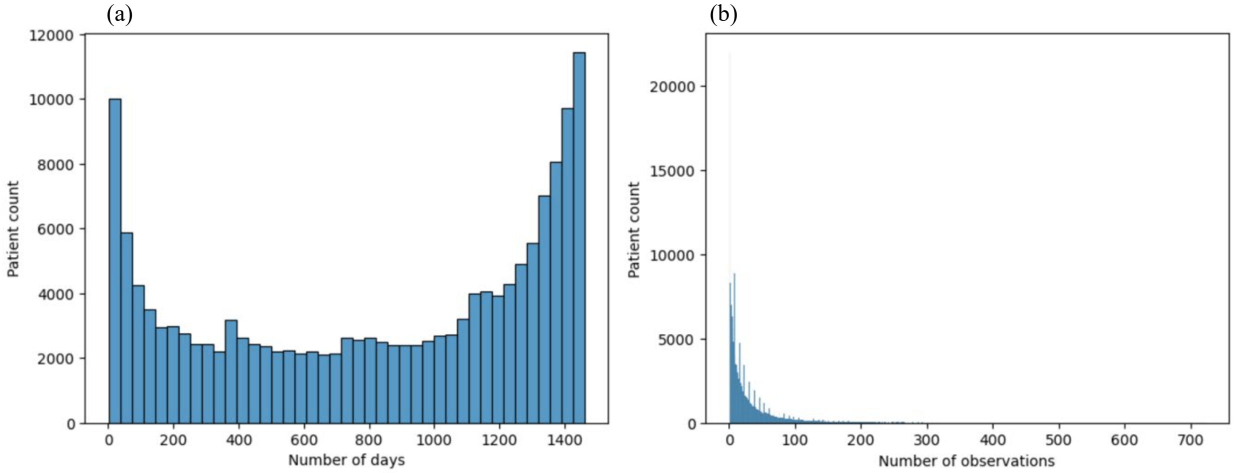


**Supplementary Figure 2 |** (a) Distribution of the length of the patient trajectories (in days). (b) Distribution of number of observations (in each patient trajectory).

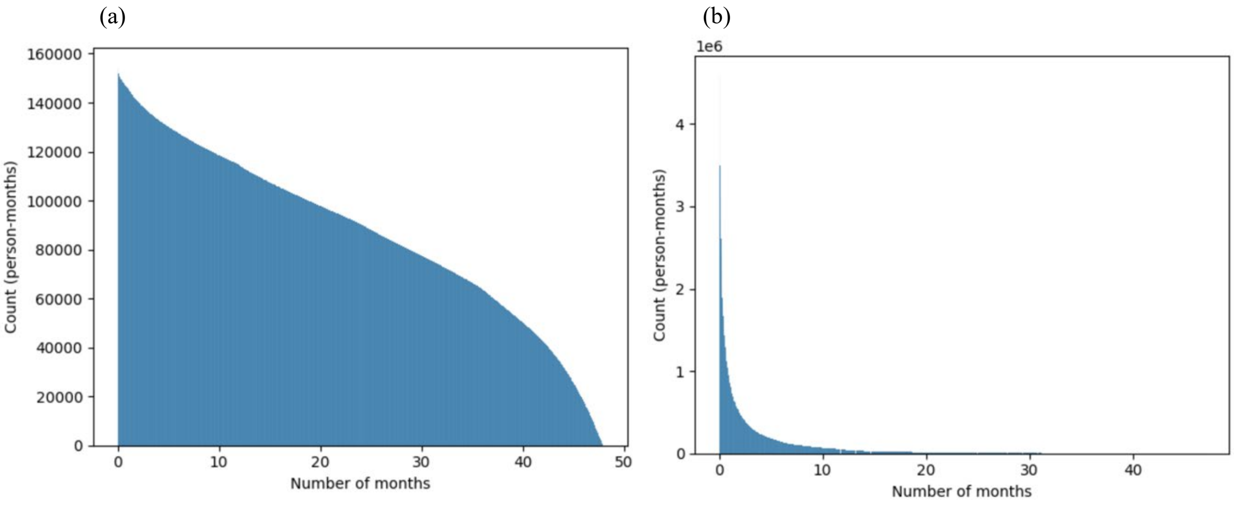


**Supplementary Figure 3 |** (a) Distribution of number of months since the first observation. (b) Distribution of number of months since each patient's previous observation in the data set.

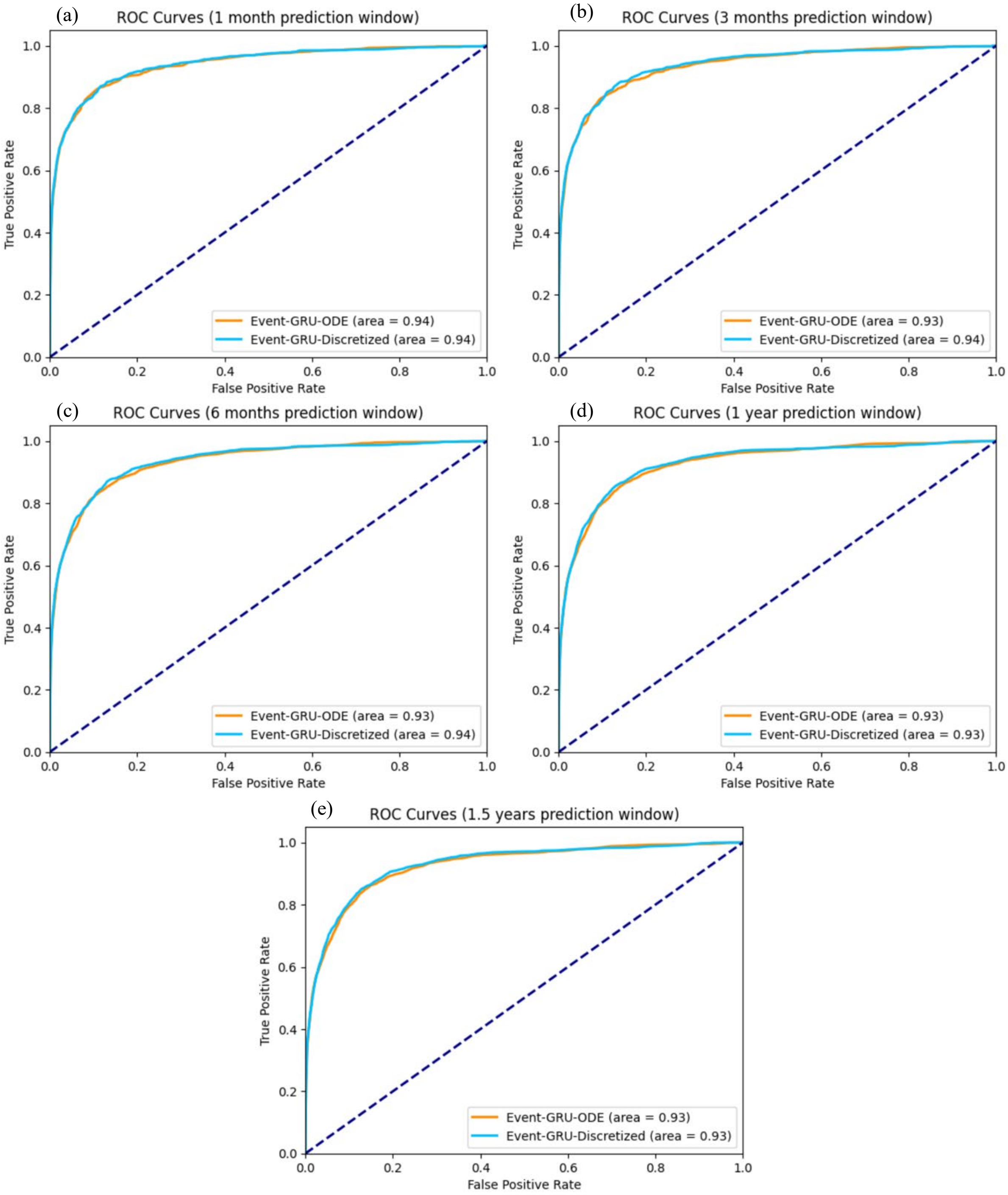


**Supplementary Figure 4 |** Receiver Operating Characteristic (ROC) curve comparison between Event-GRU-ODE and Event-GRU-Discretized, for each prediction window ((a): 1 month; (b) 3 months; (c) 6 months; (d) 1 year; (e) 1.5 years). The plots show almost identical curves obtained from the two models.

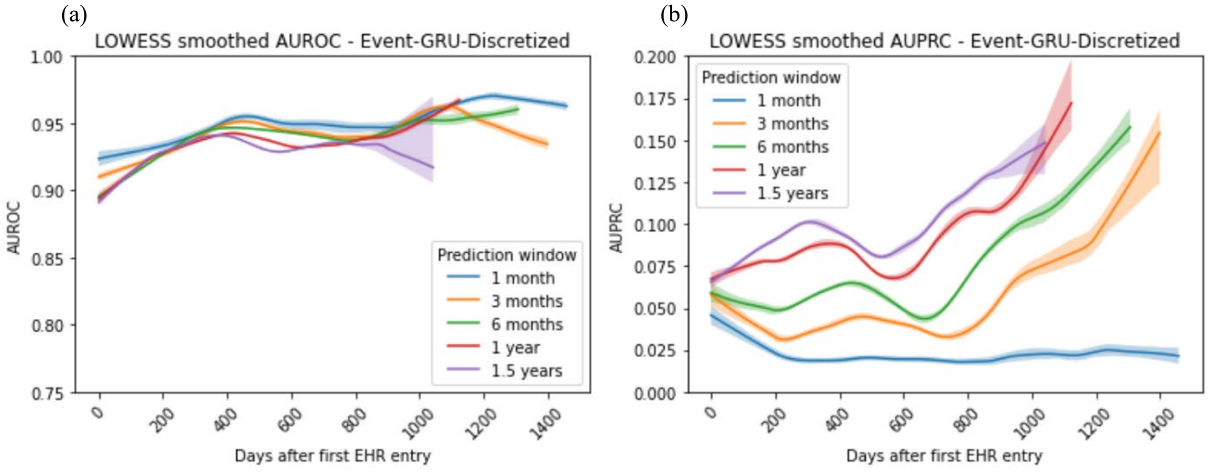


**Supplementary Figure 5 |** Aggregated prediction performance ((a) AUROC; (b) AUPRC) over time (smoothed by LOWESS) since the beginning of every patient trajectory, for Event-GRU-Discretized.

(a) (b)

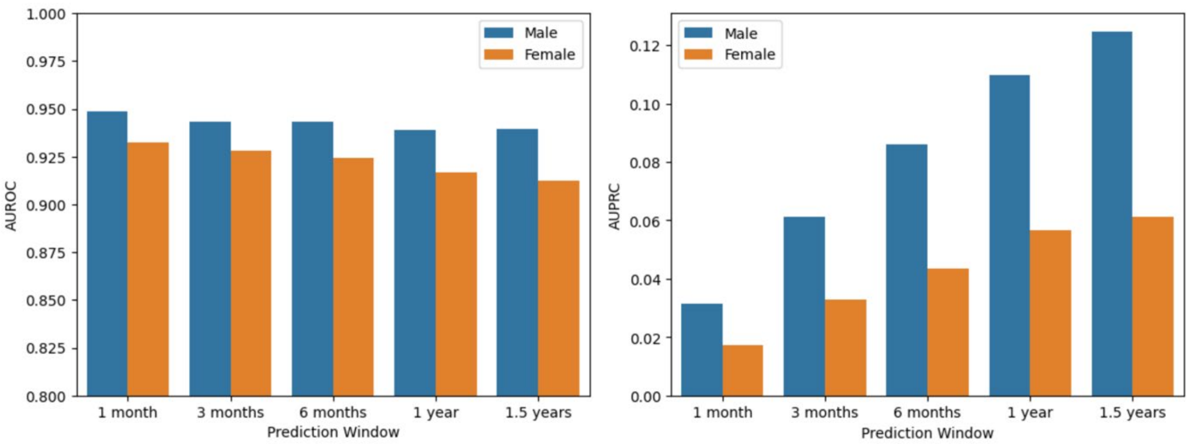


**Supplementary Figure 6 |** Model performance ((a) AUROC and (b) AUPRC) stratified by gender for Event-GRU-ODE.

(a) (b)

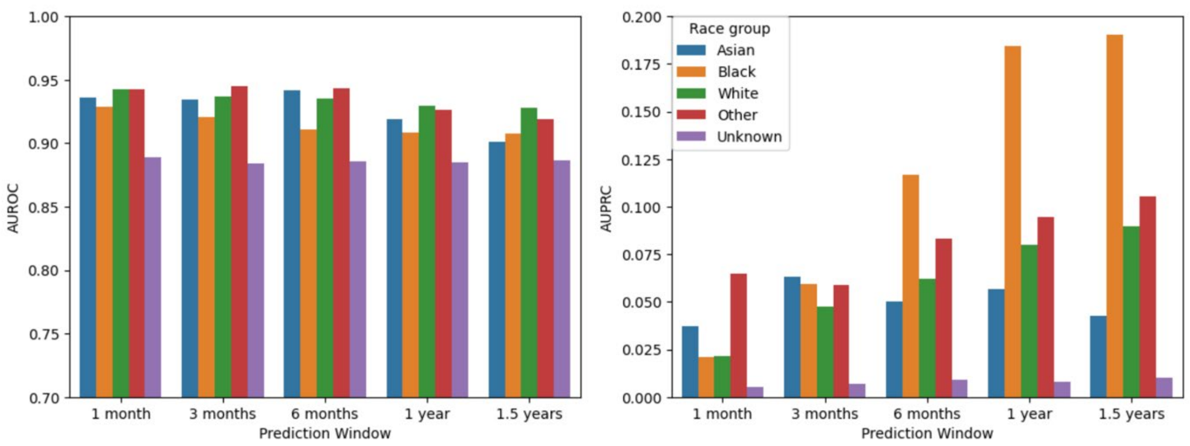


**Supplementary Figure 7 |** Model performance ((a) AUROC and (b) AUPRC) stratified by self-reported race for Event-GRU-ODE.

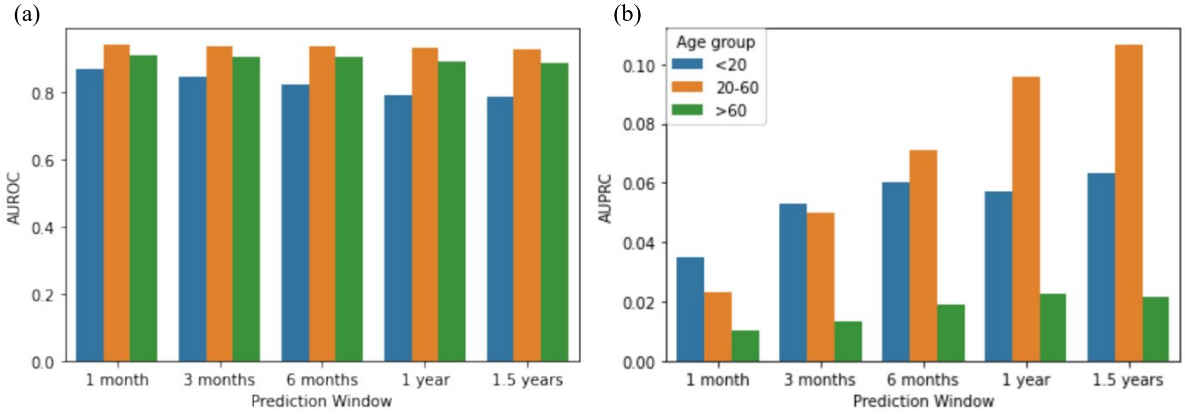


**Supplementary Figure 8 |** Model performance ((a) AUROC and (b) AUPRC) stratified by age groups for Event-GRU-ODE.

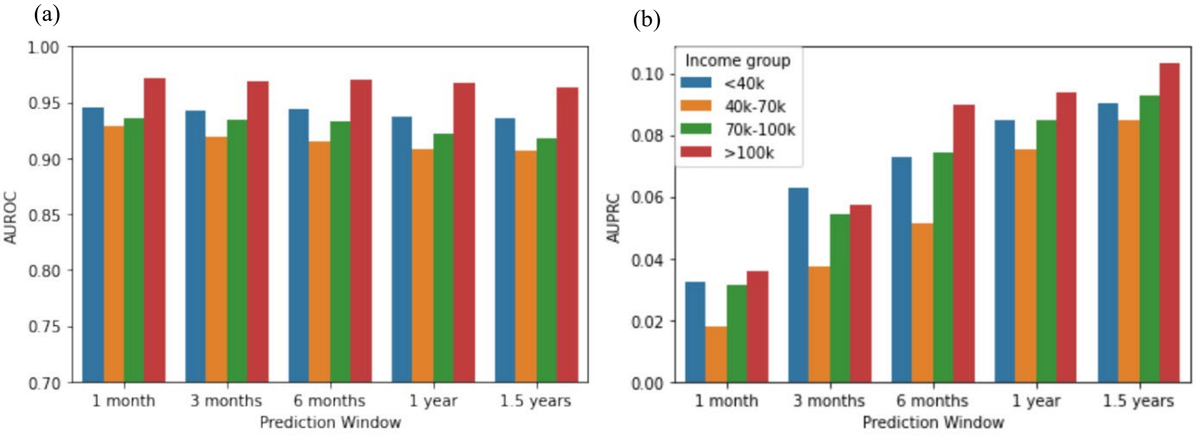


**Supplementary Figure 9 |** Model performance ((a) AUROC; (b) AUPRC) stratified by income level by ZIP codes for Event-GRU-ODE.

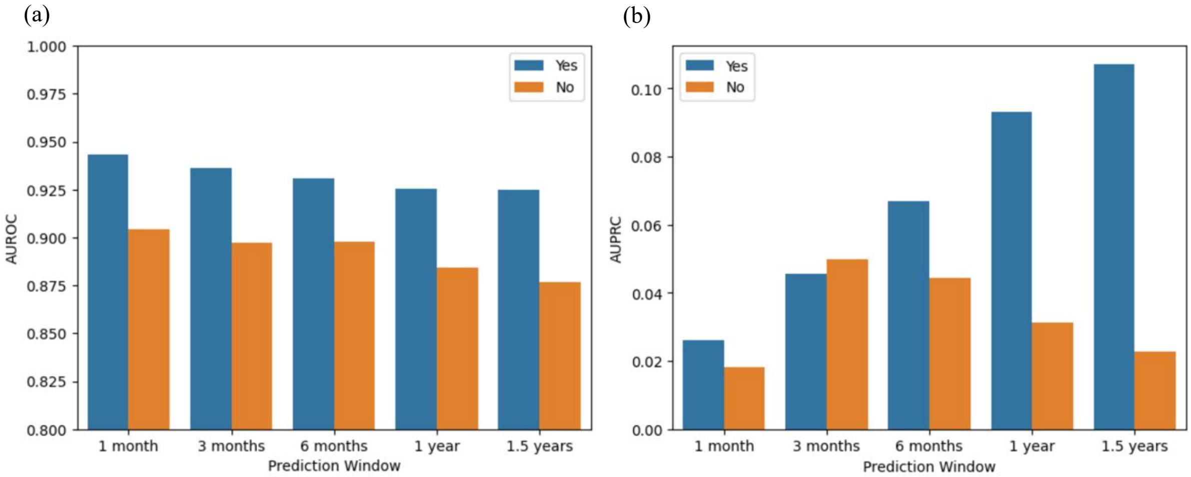


**Supplementary Figure 10 |** Model performance ((a) AUROC and (b) AUPRC) stratified by whether patients have a public payor for Event-GRU-ODE.

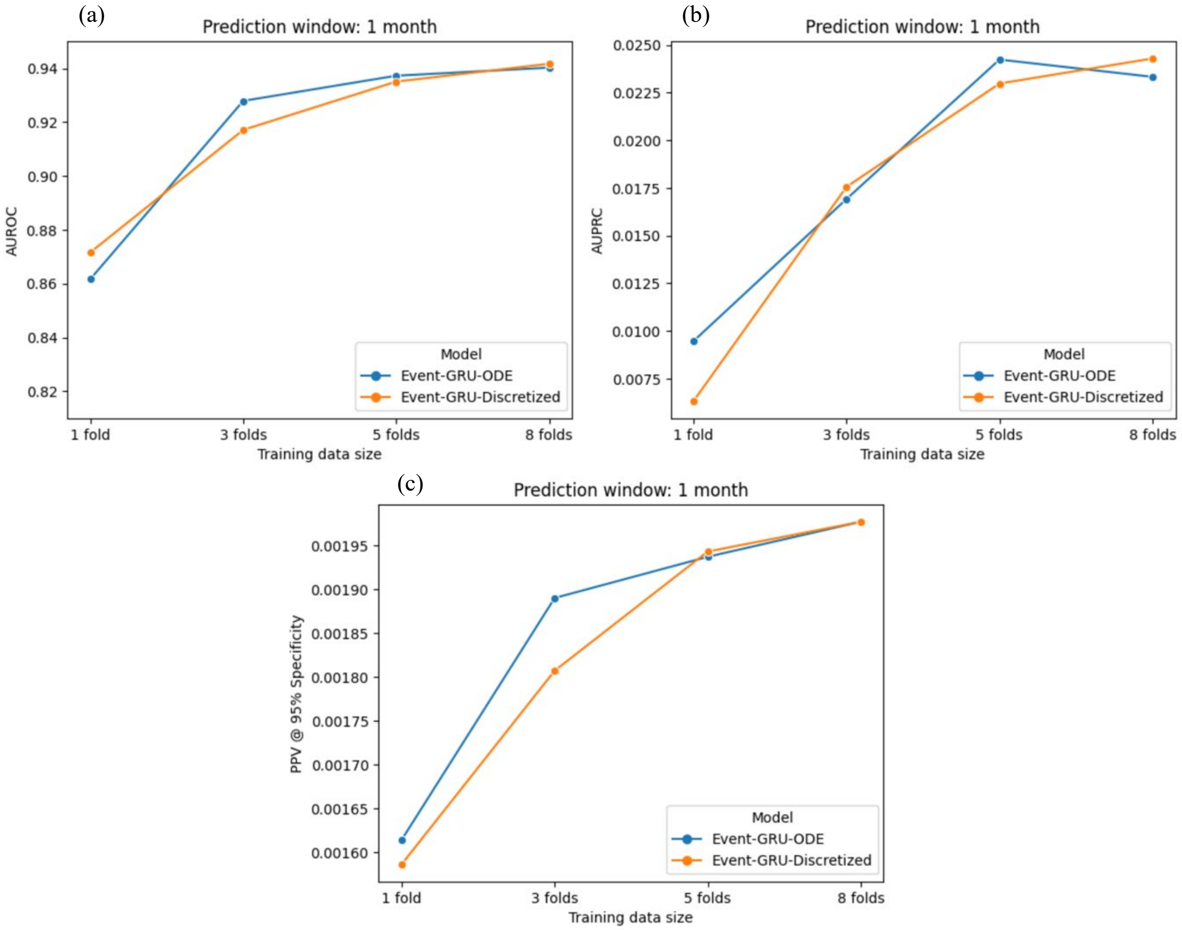


**Supplementary Figure 11 |** Model performance comparison ((a) AUROC; (b) AUPRC; (c) PPV at 95% specificity) between the two models trained with different proportions of the data set (1 fold, 3 folds, 5 folds and 8 folds), using a 1-month prediction window.

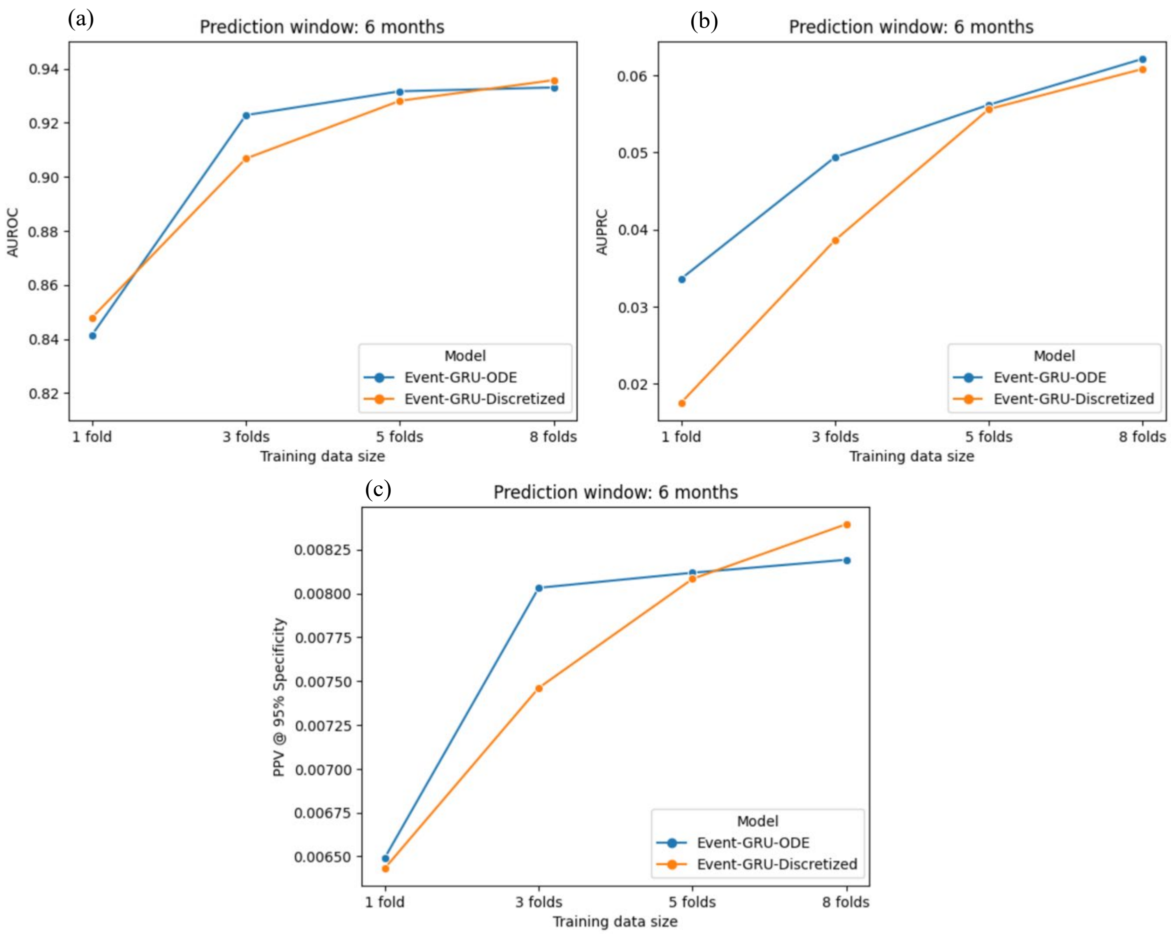


**Supplementary Figure 12 |** Model performance comparison ((a) AUROC; (b) AUPRC; (c) PPV at 95% specificity) between the two models trained with different proportions of the data set (1 fold, 3 folds, 5 folds and 8 folds), using a 6-month prediction window.

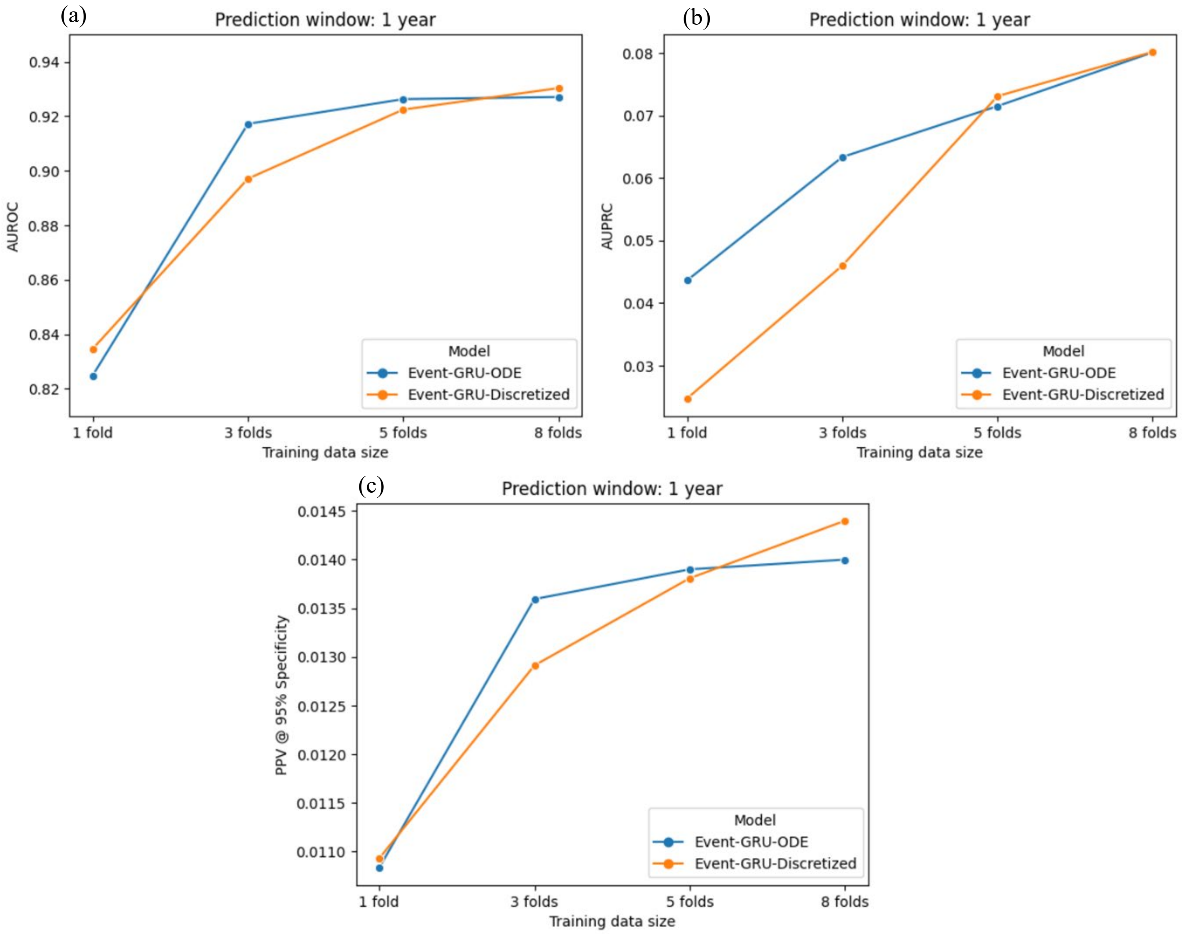


**Supplementary Figure 13 |** Model performance comparison ((a) AUROC; (b) AUPRC; (c) PPV at 95% specificity) between the two models trained with different proportions of the data set (1 fold, 3 folds, 5 folds and 8 folds), using a 1-year prediction window.

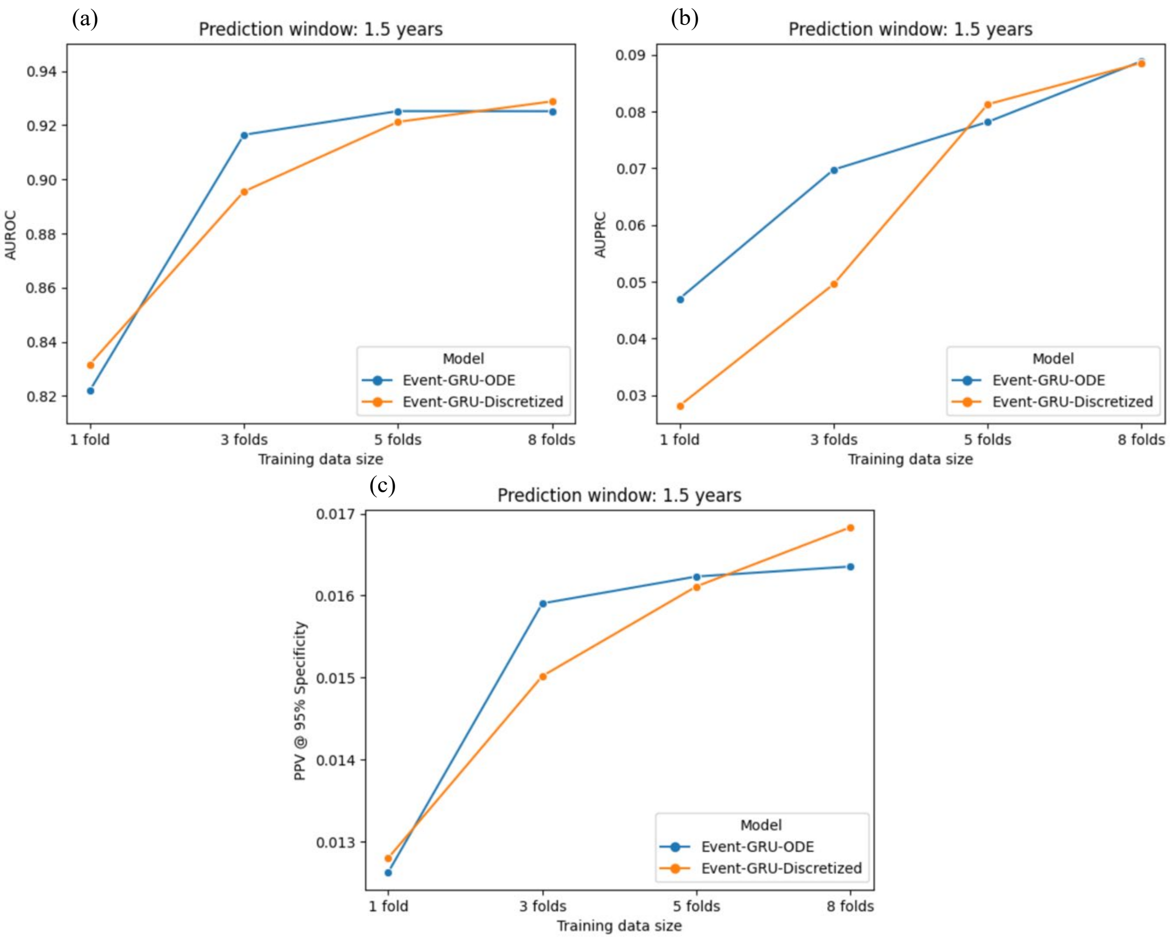


**Supplementary Figure 14 |** Model performance comparison ((a) AUROC; (b) AUPRC; (c) PPV at 95% specificity) between the two models trained with different proportions of the data set (1 fold, 3 folds, 5 folds and 8 folds), using a 1.5-year prediction window.

**Supplementary Note**

**Supplementary Note A |** Model implementation details

**Supplementary Note A |** Model implementation details

Both of the Event-GRU-ODE and Event-GRU-Discretized models are implemented using *PyTorch*. Hyperparameter selection is based on the validation set, and the tuning is performed using *HyperbandPruner* from the Python *Optuna*[35] package, with 10 trials for each model. Using validation set we optimize the hyperparameters to maximize the precision at false positive rate of 0.01 (or specificity of 0.99) after a fixed number of epochs (6 epochs for Event-GRU-ODE, and 7 epochs for Event-GRU-Discretized. Number of epochs were chosen based on preliminary testing on the training set). All models were trained and tested on a GPU cluster with 5 nodes of NVIDIA DGX-1, each consisting of 8 NVIDIA Tesla V100 GPUs with an aggregate of 1280 GB of GPU memory. For the purpose of model tuning and training, we requested for compute sessions with two and one V100 GPUs (i.e., 64 and 32 GB of GPU memory), respectively.

The best hyperparameters for the two models are:

| Hyperparameter | Event-GRU-ODE | Event-GRU-Discretized |
|---|---|---|
| **Training epochs** | 6 | 7 |
| **Learning rate** | 0.001 | 0.001 |
| **Batch size** | 200 | 200 |
| **Hidden size** | 256 | 256 |
| **Cov hidden size** | 256 | 256 |
| **Output hidden size** | 256 | 256 |
| **Cov dropout rate** | 0.2 | 0.05 |
| **Output dropout rate** | 0.2 | 0.0 |
| **Weight decay** | 4.94e-06 | 2.91e-06 |
| **ODE slower** | 10.0 | N/A |

Description of the hyperparameters:

| Hyperparameter | Description |
|---|---|
| **Training epochs** | Number of full iterations through the training data. |
| **Learning rate** | Learning rate used in the Adam optimizer. |
| **Batch size** | Number of patient trajectories used at each learning step. |
| **Hidden size** | Size of the GRU-ODE layer or GRU-Discretized layer. |
| **Cov hidden size** | Size for the hidden layer in the covariate input network. |
| **Output hidden size** | Size of the output layers. |
| **Cov dropout rate** | Dropout probability in the covariate input network. |
| **Output dropout rate** | Dropout probability in the output network. |
| **Weight decay** | Weight decay regularization used in the Adam optimizer |
| **ODE slower** | Time unit of the GRU-ODE, i.e. 10.0 means that one unit of internal clock of time in the GRU-ODE corresponds to 10 days of real time steps. Thus, the higher the value of "ODE slower" the less distance the hidden state of the GRU-ODE can move between two time steps. |
